# GWAS of amiodarone-induced thyroid dysfunction: Applications for genotype-guided risk stratification

**DOI:** 10.64898/2026.03.02.26347413

**Authors:** Søren A. Rand, Johan Bundgaard, Vinicius Tragante, Solvi Rognvaldsson, Gustav Ahlberg, Aeron M. Small, Whitney Hornsby, Satoshi Koyama, Michael Schwinn, Mart Kals, Maris Teder-Laving, Andres Metspalu, Christian Erikstrup, Mie T. Bruun, Bitten Aagard, Henrik Ullum, Søren Brunak, DBDS Genomic Consortium, Estonian Biobank Research Team, Sisse R. Ostrowski, Nanna Brøns, Jacob Træholt, Christina Mikkelsen, Bertram D. Kjerulff, Ole B. Pedersen, Erik Sørensen, Stefan Stender, Gisli Halldorsson, Ingileif Jonsdottir, Egil Ferkingstad, Hannes Helgason, Saedis Saevarsdottir, Pradeep Natarajan, Anna Helgadottir, Daniel Gudbjartsson, Henning Bundgaard, Jonas Ghouse

## Abstract

**Background:** Amiodarone is a widely used antiarrhythmic which frequently induces thyroid dysfunction, including both amiodarone-induced hypothyroidism (AIH) and thyrotoxicosis (AIT). Whether genetic factors contribute to these adverse drug reactions is unknown. In this study, we aimed to identify genetic variants that influence the risk of amiodarone-induced thyroid dysfunction and to evaluate their potential to support genotype-guided risk screening.

**Methods:** This pharmacogenetic study comprised two genome-wide meta-analyses of AIH and AIT using five datasets (Copenhagen Hospital Biobank, The Danish Blood Donor Study, Estonian Biobank, deCODE genetics, and Mass General Brigham Biobank). Key measures included the odds ratio (OR) per risk allele, the variants’ effects on spontaneous thyroid disease and biomarkers, and their clinical predictive ability, assessed by the area under the receiver operating curve (AUC), positive and negative predictive values (PPV and NPV).

**Findings:** The AIH meta-analysis (880 cases, 4,031 controls) identified three genome-wide significant loci in: *FOXE1* (rs36052460; OR 2.58, allele frequency [AF] = 64.3%, *P* = 2.55 × 10^−44^), *FOXA2* (rs2424459; OR 1.67, AF = 71.3%, *P* = 2.59 × 10^−14^), and *ADAM32* (rs12681571; OR 1.49, AF = 61.8%, P = 3.05 × 10^−9^). The AIT meta-analysis (385 cases, 4,936 controls) identified one locus in *CAPZB* (rs867355; OR 1.63, AF = 66.1%, *P* = 3.49 × 10^−8^). In risk prediction models, a polygenic risk score (PRS) of the AIH variants increased the AUC by 9.2% (95% CI 6.6 – 11.9%), which outperformed a genome-wide hypothyroidism PRS (1.5% AUC increase, 95%CI 0.0 – 2.9%). Similarly, the *CAPZB* variant improved AIT prediction (AUC increase of 4.0%, 95% CI 0.4 – 7.5%) beyond a hyperthyroidism PRS (0.2% AUC increase, 95%CI -0.8 – 1.2%). Genotype-guided screening would identify individuals at low risk (NPVs ranging from 90-95% and PPVs 2-20%).

**Interpretation:** We identified genetic variants that influence the risk of developing amiodarone-induced thyroid dysfunction. Genotype-guided screening offers a potential complement to current strategies and personalize pre-treatment risk assessment for patients initiating amiodarone therapy.

## INTRODUCTION

Amiodarone is a widely used potent antiarrhythmic effective for managing atrial fibrillation and preventing ventricular arrhythmias.^1,2^ However, its therapeutic benefits are often offset by a range of both acute and insidious adverse drug reactions (ADRs). While these frequently affect the eyes, skin, liver, and lungs, thyroid dysfunction is one of the most common and clinically challenging.^1^

Amiodarone-induced thyroid dysfunction manifests as either hypothyroidism (AIH) or thyrotoxicosis (AIT). AIH, which affects 10–20% of patients, typically develops within six months of therapy and is associated with female sex, elevated thyroid stimulating hormone (TSH), and positive anti-thyroid peroxidase (anti-TPO) antibodies.^3^ AIT affects 5–10% of patients, usually within one year of therapy, with risk factors including male sex, iodine-deficiency, and positive thyrotropin-receptor antibodies (TRAb).^2,4^ Both conditions can carry significant morbidity, requiring coordinated management by both cardiologists and endocrinologists.^3–5^ AIH often presents with nonspecific symptoms (e.g. dry skin, malaise) that can delay diagnosis,^2^ and improper management can exacerbate pre-existing cardiac conditions. AIT represents a more severe clinical challenge; its thyrotoxic component can precipitate angina, trigger arrythmias, or worsen left ventricular systolic dysfunction.^2^ In high-risk patients, such as those with heart failure, AIT mortality can range from 30% to 50%, and, in severe cases, trigger a life-threatening thyroid storm.^2,3^

The current strategy to mitigate thyroid ADRs is reactive, relying on labor-intensive biochemical monitoring rather than predicting individual-level risk. Genome-wide association studies (GWAS) have identified numerous variants linked to spontaneous thyroid disease and function, underscoring the strong genetic basis of thyroid regulation.^6–10^ However, it remains unknown whether the same genetic architecture influences the risk of amiodarone-induced thyroid dysfunction. In this pharmacogenetic study, we interrogated the genetic basis of AIH and AIT to identify variants that could support genotype-guided risk assessment and improve pre-therapy safety evaluation.

## METHODS

### Datasets

We analyzed five datasets: The Copenhagen Hospital Biobank (CHB), The Danish Blood Donor Study (DBDS), deCODE genetics, The Estonian Biobank (EstBB), and Mass General Brigham Biobank (MGBB). CHB comprises hospital patients sampled during admissions in Denmark’s Capital Region, while DBDS includes voluntary blood donors recruited nationwide, together totaling over 380,000 genotyped individuals.^11,12^ deCODE genetics includes genomic data on approximately half of the Icelandic population, linked with extensive phenotypic information.^13,14^ The EstBB is a population-based study of over 200,000 individuals representative of the adult Estonian population by age, sex and geographic distribution.^15,16^ The MGBB is a hospital-based biobank containing data from over 65,000 genotyped patients from Massachusetts General Hospital and Brigham and Women’s Hospital (**Supplementary Table 1; Supplementary Materials**).^17^ Genotyping was performed on various Illumina platforms. We applied standard sample- and variant-level quality control, retaining only well-genotyped, common single nucleotide polymorphisms (SNPs) and removing individuals with sex discordance, excessive missingness, or non-European ancestry. Quality control and imputation procedures for each dataset are described in **Supplementary Table 1**.

### Samples and study design

We first identified all individuals prescribed amiodarone (Anatomical Therapeutic Chemical [ATC] code: C01BD01). Using established timelines for ADR onset,^4^ we defined cases and controls for AIH and AIT as described below:

- AIH cases: Defined as individuals who received a hypothyroidism diagnosis (ICD-10: E03[2, 8, or 9]) or a prescription for thyroid hormone therapy (ATC: H03A) within 365 days of their first amiodarone prescription. Individuals with any history of hypothyroidism or thyroid hormone use prior to amiodarone initiation were excluded.

AIH controls: Defined as individuals with three or more amiodarone prescriptions within one year who had no history of hypothyroidism or thyroid hormone use.

- AIT cases: Defined as individuals who received a thyrotoxicosis diagnosis (ICD-10: E05[8 or 9]) or a prescription for antithyroid medication (ATC: H03B) within 550 days of their first amiodarone prescription. Those with prior diagnoses or treatment were excluded.

AIT controls: Defined as individuals with three or more amiodarone prescriptions within a year and no diagnosis of hyperthyroidism or use of antithyroid medication.

### Association testing, quality control and meta-analysis

In CHB, DBDS, and deCODE genetics we used logistic regression, adjusting for age at first prescription, sex, and population stratification (4 principal components [PCs] in CHB and DBDS and county of origin for deCODE genetics), using software developed at deCODE genetics. In Estonian Biobank and MGBB, we used REGENIE,^18^ adjusting for age at first prescription, sex, and 10 PCs. Post-regression QC excluded variants with MAF < 1%, INFO < 0.7, and absolute beta > 5 and standard error > 10. We used METAL to meta-analyze all datasets employing the fixed-effect inverse-variance weighted method,^19^ and the LD Score Regression (LDSC) to calculate SNP heritability and genomic inflation using linkage disequilibrium (LD) scores calculated in the HapMap3 CEU reference panel.^20^ We set traditional genome-wide significance (*P* < 5 × 10^−8^) as criteria for significance, and at each locus defined lead SNPs as the genetic variant with the lowest *P*-value.

### Comparison of genetic architecture

To compare the genetic architecture of amiodarone-induced thyroid dysfunction to spontaneous thyroid disease, we performed a series of cross-trait analyses. First, we identified independent, genome-wide significant SNPs for spontaneous hypothyroidism from the largest available GWAS, and for hyperthyroidism, we queried variants reported in the GWAS-catalogue.^8,9,21,22^ We then extracted β-coefficients for these lead variants, plotted them against their corresponding effects on AIH and AIT, and assessed correlations using Pearson’s test. Second, we formally tested for shared causal signals at overlapping loci using coloc.^23^ A posterior probability for H4 (PP4) > 0.8 was considered strong evidence for a shared signal, while a PP3 > 0.8 suggested distinct signals. Finally, we evaluated our primary ADR-associated variants for their effects on spontaneous hypothyroidism and hyperthyroidism, and thyroid hormone levels ([TSH], thyroglobulin [Tg], free thyroxine [fT4]). We extracted these effect estimates from the largest available publications,^9,22,24^ and used Cochran’s Q test to identify significant heterogeneity (*P* _*Het*_ < 0.05) between the variant effects on amiodarone-induced versus spontaneous disease.

### Gene mapping and characterization

We prioritized and functionally characterized candidate genes in amiodarone-induced thyroid diseases using the following four methods:

1. Distance to nearest protein coding gene.
2. Protein altering variants that were within a ± 250 Kb window of the lead variant and in high linkage disequilibrium (LD; *r*^2^ > 0.8).
3. Polygenic Priority Score (PoPS): PoPS is a similarity-based gene prioritization tool that integrates GWAS summary statistics with functional genomic data, including biological pathways, chromatin accessibility, gene expression, and protein-protein interactions.^25^ PoPS calculates an *in-silico* score (PoPS-score) for each gene, which represents the likelihood of the gene being associated to the trait. We considered genes in a ± 250 kb window around the lead variant and prioritized the gene with the highest PoPS-score.
4. Expression quantitative trait loci (eQTL): We used all tissues from Genotype-Tissue Expression (GTEx)-v10,^26^ and only considered *cis*-eQTL associations (within 1 Mb of lead variant positions). In each locus, we assessed both the effect on RNA expression from the lead variants and the top eQTL signals in the locus and also considered LD between lead variants and top eQTL signals.

### Risk prediction models

In CHB and DBDS, we evaluated the predictive performance of five risk factors: Age, sex, polygenic risk scores (PRS), AIH/AIT-associated variants, and thyroid autoantibodies. We first constructed a benchmark model including age, sex, first four PCs, and pre-existing thyroid disease. We then expanded the model by adding risk factors iteratively with performance assessed in three settings:

i. In AIH/AIT.
ii. In AIH/AIT individuals with thyroid autoantibodies (anti-TPO for AIH or TRAb for AIT).
iii. In spontaneous disease (hypothyroidism or hyperthyroidism), to evaluate if the risk factors also predicted spontaneous disease forms.

Detailed definitions of each the above are provided in **Supplementary Table 2**. Model performance was assessed by comparing the change in area under the curve receiver operating characteristic (AUC) relative to the benchmark model using the R-package pROC.^27^ Significant differences between models were tested using DeLong’s test for correlated ROC curves. We constructed a PRS_AIH_, by weighting the effect size of each AIH-associated variant’s risk allele, summing, and then scaling the score to a mean of zero and a standard deviation of 1. PRSs for hypothyroidism and hyperthyroidism were computed based on published genome-wide PRS-weights derived from UK Biobank, available through the PGS-catalog: PRS_hypo_, N_case_: 16,945, N_control_: 371,311, URL: https://www.pgscatalog.org/score/PGS002024/, PRS_hyper_, N_case_: 2,103, N_control_: 371,311, URL: https://www.pgscatalog.org/score/PGS002023/.

### Clinical validity and utility of screening for ADR-associated variants

We assessed the clinical utility of genotype-based screening for ADR-associated variants by calculating the sensitivity, specificity, positive and negative predictive values (PPV, NPV), and numbers needed to genotype (NNG).^28^ We used PLINK^29^ to extract individual genotypes of the ADR-associated variants and also included the PRS_AIH_ for evaluation. Analyses were done in CHB/DBDS, deCODE genetics, and the UK Biobank primary care dataset (external validation resource including data from approximately 245,000 participants available for non-COVID-related research). We applied the same case-control definitions as in the association analyses, and evaluated screening performance as described below:

1. AIH and *FOXE1:* We used positive tests as carrying either one or two risk alleles.
2. AIH and *FOXA2:* We used positive tests as carrying either one or two risk alleles.
3. AIH and *ADAM32:* We used positive tests as carrying either one or two risk alleles.
4. AIH and PRS _AIH_ : We used PRS cut-offs at both top and bottom 10, 20, and 30^th^ deciles as positive tests.
5. AIT and *CAPZB:* We used positive tests as carrying either one or two risk alleles.

## RESULTS

### Clinical characteristics of patients with amiodarone-induced thyroid dysfunction

Across all five datasets, we identified 880 AIH cases and 4,031 amiodarone-treated controls without hypothyroidism (patient demographics in **Supplementary Table 3**). Consistent with prior reports, females were more likely to develop AIH.^4^ We found no consistent differences in comorbidities or use of other antiarrhythmic medication between cases and controls. For AIT, we found 385 cases and 4,936 amiodarone-treated controls without hyperthyroidism (patient demographics in **Supplementary Table 4**). Similar to AIH, there were no consistent differences in comorbidities or use of anti-arrhythmic medication.

### Genetic architecture of amiodarone-induced thyroid dysfunction

In the GWAS meta-analysis of AIH, we identified three genome-wide significant loci (*P* < 5 × 10^−8^; **Table 1; Figure 1A**). The strongest signal was observed for rs36052460, located in an enhancer site 67 Kb downstream of the thyroid transcription factor *FOXE1* (OR 2.58, 95% CI 2.26 - 2.95, *P* = 2.55 × 10^−44^, risk allele frequency [RAF] = 64.3%),^30^ and was previously associated with hypothyroidism, autoimmune thyroid disease, thyroid carcinoma, free thyroxine (fT4), and thyroid stimulating hormone (TSH) levels.^31–33^ The next AIH-associated variant, rs2424459 near *FOXA2* (OR 1.67, 95%CI 1.47 - 1.91, *P* = 2.59 × 10^−14^, RAF = 64.3%), also had prior association with hypothyroidism and TSH.^31^ The final lead variant rs12681571, near *ADAM32* (OR 1.49, 95% CI 1.31 - 1.75, *P* = 3.05 × 10^−9^, RAF = 61.8%), had no previous associations with other traits. For AIT, we identified one genome-wide significant variant, rs867355 near *CAPZB* (OR 1.63, 95% CI 1.37 – 1.94, *P* = 3.05 × 10^−8^, RAF = 66.1%) which was previously associated with hypothyroidism, fT4, and TSH.^31^ Heritability analyses were not performed due to low statistical power (**Supplementary Table 5**). Regional association plots are shown in **Supplementary Figure 1** and per dataset summary data in **Supplementary Table 6**.

**Table 1.** Genome-wide independent associations for amiodarone-induced hypothyroidism (AIH) and thyrotoxicosis (AIT). Results from a meta-analysis of AIH (880 cases and 4,031 controls) and AIT (385 cases and 4,936 controls) using five European ancestry biobanks (Copenhagen Hospital Biobank, Danish Blood Donor Study, deCODE genetics, Estonian Biobank, and Mass General Brigham Biobank). *P*-values were two-sided and based on an IVW fixed-effects meta-analysis. Abbreviations: AIT/AIH, amiodarone-induced thyrotoxicosis/hypothyroidism; RA/OA risk allele and other allele; RAF, risk allele frequency; OR, odds ratio; CI, confidence interval; PP3, the posterior probability for the association with both traits is driven by different causal variants; PP4, the posterior probability for the association with both traits is driven by a single shared causal variant. Genomic positions are according to GRCh38.

| Trait | SNP | Position | Nearest gene | Variant | RA/OA | RAF | OR (95%CI) | *P*-value | *I*<sup>2</sup> (%) | *P*<sub>Het</sub> | PP3 with spontaneous counterpart | PP4 with spontaneous counterpart |
| --- | --- | --- | --- | --- | --- | --- | --- | --- | --- | --- | --- | --- |
| AIH | rs12681571 | 8:39345210 | *ADAM32* | Intron | G/C | 61.8% | 1.49 (1.30 – 1.70) | $3.05 \times 10^{-9}$ | 0% | 0.77 | 97.7% | 0.1% |
| | rs36052460 | 9:97777236 | *FOXE1* | Intergenic | AT/A | 64.3% | 2.59 (2.26 – 2.95) | $2.55 \times 10^{-44}$ | 77.9% | 0.03 | 2.4% | 97.2% |
| | rs2424459 | 20:22667434 | *FOXA2* | Intron | A/C | 71.3% | 1.67 (1.47 – 1.92) | $2.59 \times 10^{-14}$ | 60.4% | 0.06 | 58.4% | 41.2% |
| AIT | rs867355 | 1:19373292 | *CAPZB* | Intron | A/G | 66.1% | 1.63 (1.37 - 1.94) | $3.05 \times 10^{-8}$ | 0% | 0.60 | 51.1% | 48.8% |

**Figure 1.**
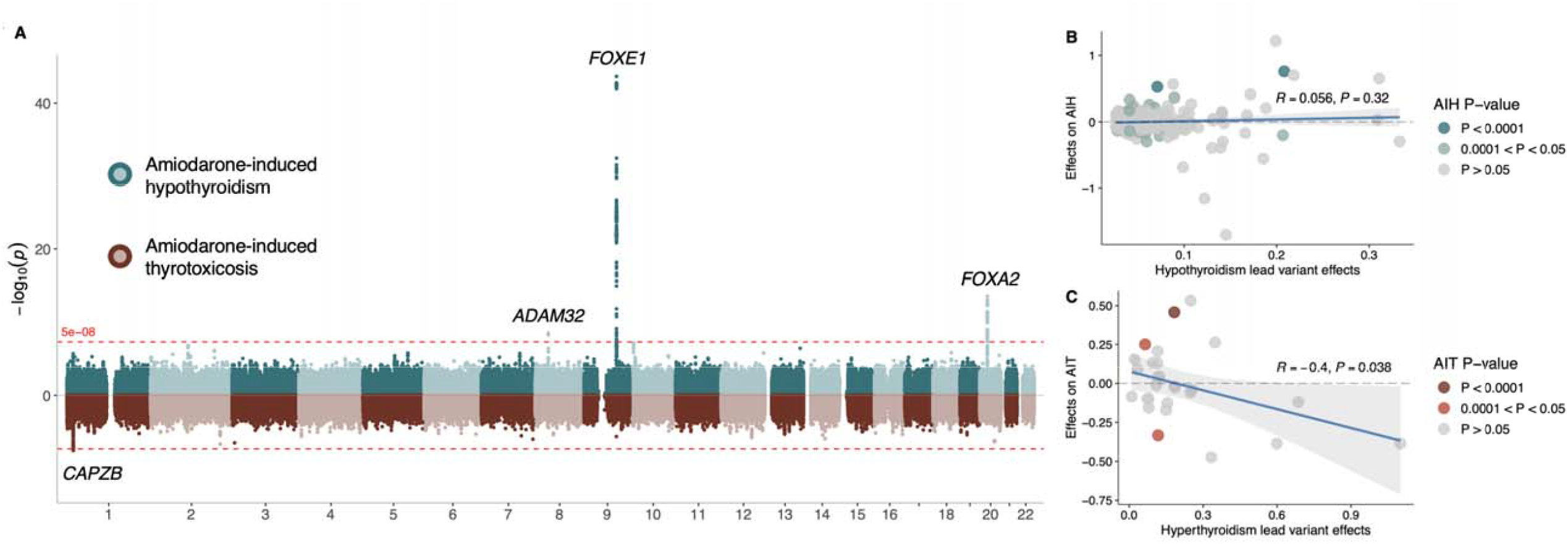
Genetic associations for amiodarone-induced hypothyroidism (AIH) and thyrotoxicosis (AIT) and their relation to counterpart spontaneous disease. A. Miami plots for amiodarone-induced hypothyroidism (top, blue) and amiodarone-induced thyrotoxicosis (bottom, red). The *x* axis is the genomic position of single nucleotide polymorphisms (SNPs), and the *y* axis is the strength of association −log_10_(*P*) between the SNP and trait. The dashed red lines indicate the threshold for genome-wide significance (*P*⍰< ⍰5⍰× ⍰10^−8^). **B and C**. We queried published GWAS data for hypothyroidism (**B**) and hyperthyroidism (**C**) and evaluated lead variants effect on their counterpart amiodarone-induced thyroid dysfunction trait. We used Pearson’s correlation to quantify strength and direction of variants relationships between traits.

In relation to these variants previous associations to spontaneous thyroid disease, we investigated whether the effects of lead variants for hypothyroidism and hyperthyroidism were comparable with those of AIH and AIT, respectively. Of 318 hypothyroidism lead variants, 303 were available in the AIH dataset; we found no significant correlation in effects between hypothyroidism and AIH (Pearson’s *r* = 0.06, *P* = 0.32; **Figure 1B**), and only the *FOXE1* locus colocalized between hypothyroidism and AIH (PP4 and PP3 values shown in **Table 1**). Next, we assessed 27 independent hyperthyroidism lead variants effects on AIT and observed a negative correlation (Pearson’s *r* = -0.40, *P* = 0.039; **Figure 1C**), and no evidence of colocalization at the *CAPZB* locus (**Table 1**; colocalization plots in **Supplementary Figure 2A–C**).

### Characterization of genetic effects on thyroid disease and function

We next focused on the effects of the ADR-associated variants on hypothyroidism, hyperthyroidism, fT4, TSH, and Tg levels (**Supplementary Table 7; Figure 2**). The *FOXE1* variant rs36052460, which colocalized between AIH and hypothyroidism, conferred a significantly higher risk of AIH relative to spontaneous hypothyroidism (OR_AIH_ 2.58 vs. OR_Hypo_ 1.27, *P*_*Het*_ = 2.11 × 10^−25^), indicating a possible gene-environment interaction with amiodarone. The risk allele was also associated with increased TSH and fT4 levels. The *FOXA2* variant rs2424459 also showed significantly larger effect on AIH than on spontaneous hypothyroidism (OR_AIH_ 1.67 vs. OR_Hypo_ 1.05, *P*_*Het*_ = 7.43 × 10^−12^) and was associated with increased TSH levels, slightly reduced fT4 levels, and a large decrease in Tg. The *ADAM32* variant rs12681571, which had no previous genome-wide associations, exhibited a weak, nominally significant opposing effect on spontaneous hypothyroidism (OR_AIH_ 1.49 vs. OR _Hypo_ 0.98, *P*_*Hypo*_ = 0.039, *P*_*Het*_ = 1.52 × 10^−9^) and had no effect on thyroid hormone levels. On the hyperthyroid end of the spectrum, the *CAPZB* variant rs867355 associated with higher risk of AIT compared with that of spontaneous hyperthyroidism (OR_AIT_ 1.63 vs. OR_Hyper_ 1.09, *P*_*Het*_ = 5.84 × 10^−6^) and was associated with suppressed TSH, and elevated Tg and fT4 levels.

**Figure 2.**
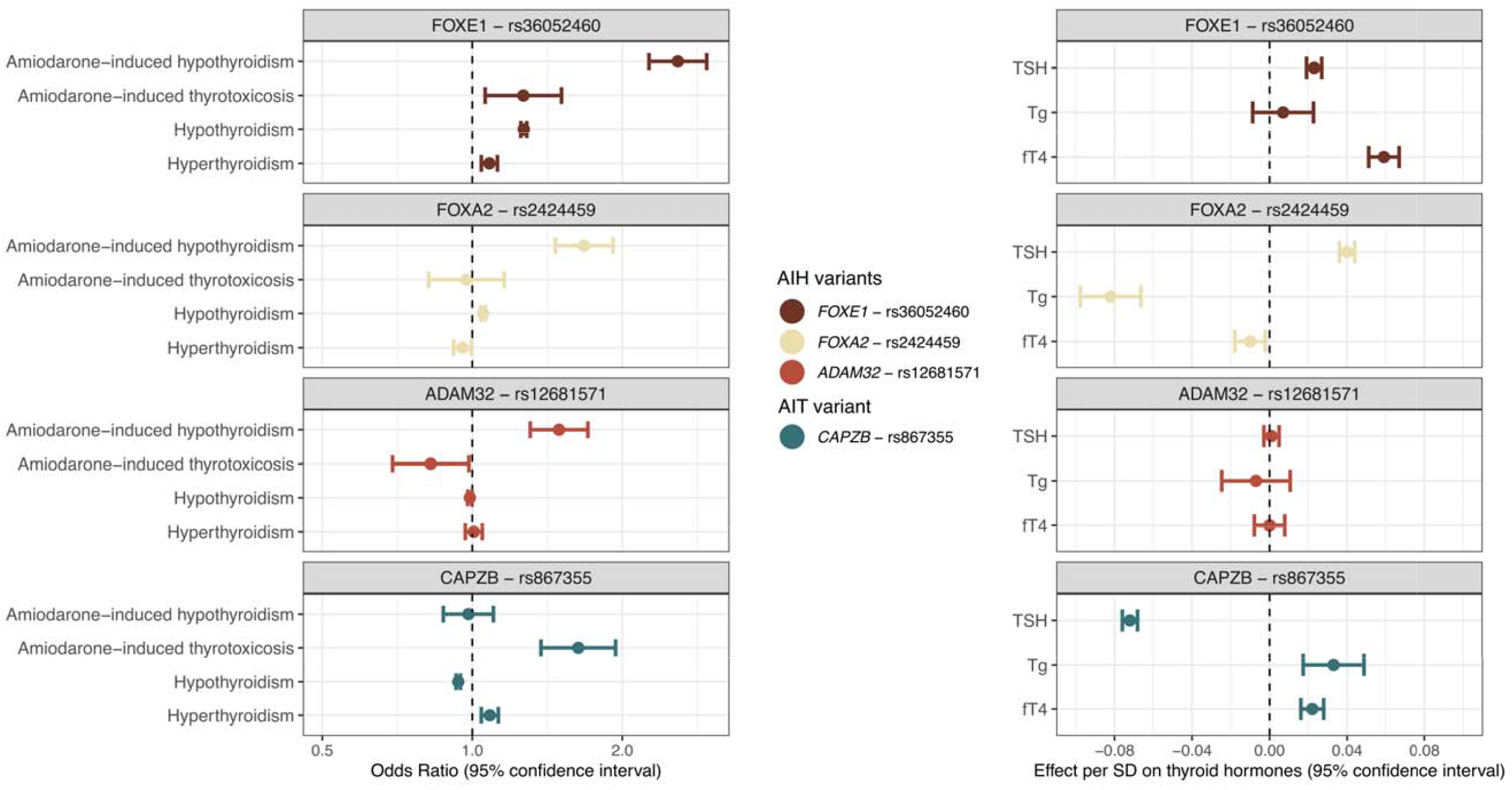
Genetic effects of lead variants in amiodarone-induced hypothyroidism (AIH) and thyrotoxicosis (AIT) on thyroid disease and hormone levels. Effects on thyroid hormone levels are presented in standardized units (SD increase). Abbreviations: AIT, amiodarone-induced thyrotoxicosis; AIH, amiodarone-induced hypothyroidism; TSH, thyroid stimulating hormone; Tg, thyroglobulin; fT4, free thyroxine.

### Gene prioritization and functional characterization

To link the identified loci to effector genes, we used four complementary approaches: (1) distance to nearest gene, (2) protein-altering coding-variant mapping, (3) top *cis-*eQTL mapping, (4) Polygenic Priority Score (PoPS). AIH/AIT variants were not in high LD (r^2^ > 0.8) with any coding variants. The AIH lead variant, rs36052460, was mapped to its closest gene, *FOXE1*, by PoPS but had no top eQTL evidence. The second AIH variant, rs2424459, was closest to *FOXA2* and in moderate LD with the top thyroid-specific *cis-* eQTL for *FOXA2* (rs1007202, r^2^ = 0.58), which was also prioritized by PoPS. The third variant, rs12681571, was mapped to its closest gene, *ADAM32*, by PoPS, but no evidence of top *cis*-eQTLs was found. The AIT variant, rs867355, was closest to *CAPZB*, and was in high LD with the top *cis*-eQTL for *SLC66A1* in hypothalamus (rs10753558, r^2^ > 0.99) and in moderate LD with the top *SLC66A1* eQTL in thyroid, (rs214313, r^2^ = 0.63). Additionally, the variant associated with thyroid expression of its PoPS prioritized gene, *CAPZB*, conditioned on the top *cis*-eQTL for *CAPZB* (rs10917451, β_unadj_ = 0.22, *P* = 4.4 × 10^−17^, β_adj_ = 0.12, *P* = 1.3 × 10^−6^), demonstrating the variant’s pleiotropic cis-effects on RNA expression (PoPS results in **Supplementary Table 8**, eQTL results in Supplementary Table 9 and Supplementary Figure 3).

### Distinct genetic predictors for amiodarone-induced and spontaneous thyroid disease

We next evaluated the predictive ability of ADR-associated variants and PRSs in individuals from CHB/DBDS in three different data samples: (1) In individuals with AIH/AIT, (2) in a subset of these with thyroid autoantibodies available, and (3) in individuals with spontaneous disease forms (sample sizes for all six data samples are shown in **Figure 3**). First, we modelled risk for amiodarone-induced thyroid dysfunction. For AIH, a benchmark model (age, sex, pre-existing thyroid disease and four PCs) achieved an AUC of 0.66. While a PRS for spontaneous hypothyroidism (PRS_hypo_) offered incremental improvement (ΔAUC_PRS-hypo_ +1.5%, 95%CI 0.0 - 2.9%), the *FOXE1* and *FOXA2* variants had much stronger impact. The best model performance was observed for a model including a PRS for AIH, increasing the AUC by +9.2% (95% CI 6.6 – 11.9%; **Figure 3A** and **Supplementary Table 10A**). For AIT, adding the PRS for spontaneous hyperthyroidism (PRS_hyper_) to the benchmark model had no significant effect (ΔAUC +0.2%, 95%CI -0.8 - 1.2%), while adding the *CAPZB* variant significantly improved risk prediction by +4.0% (95% CI 0.4 – 7.5%; **Figure 3B** and **Supplementary Table 10B**). In models including thyroid autoantibodies, variants in *FOXA1* and *FOXE2* and the PRS_AIH_ all added independent information to prediction of AIH (**Figure 3C** and **Supplementary Table 11A**). For AIT, adding the *CAPZB* variant to the TRAb-inclusive model resulted in a non-significant AUC increase of 3.5% (95% CI -3.8 – 10.8%; **Figure 4D** and **Supplementary Table 11B**). Conversely, when predicting spontaneous thyroid disease, the patterns were reversed. The PRSs were the superior predictors, outperforming the individual ADR-associated variants in models for both spontaneous hypothyroidism and hyperthyroidism (**Figure 3E-F** and **Supplementary Table 12A-B**).

**Figure 3.**
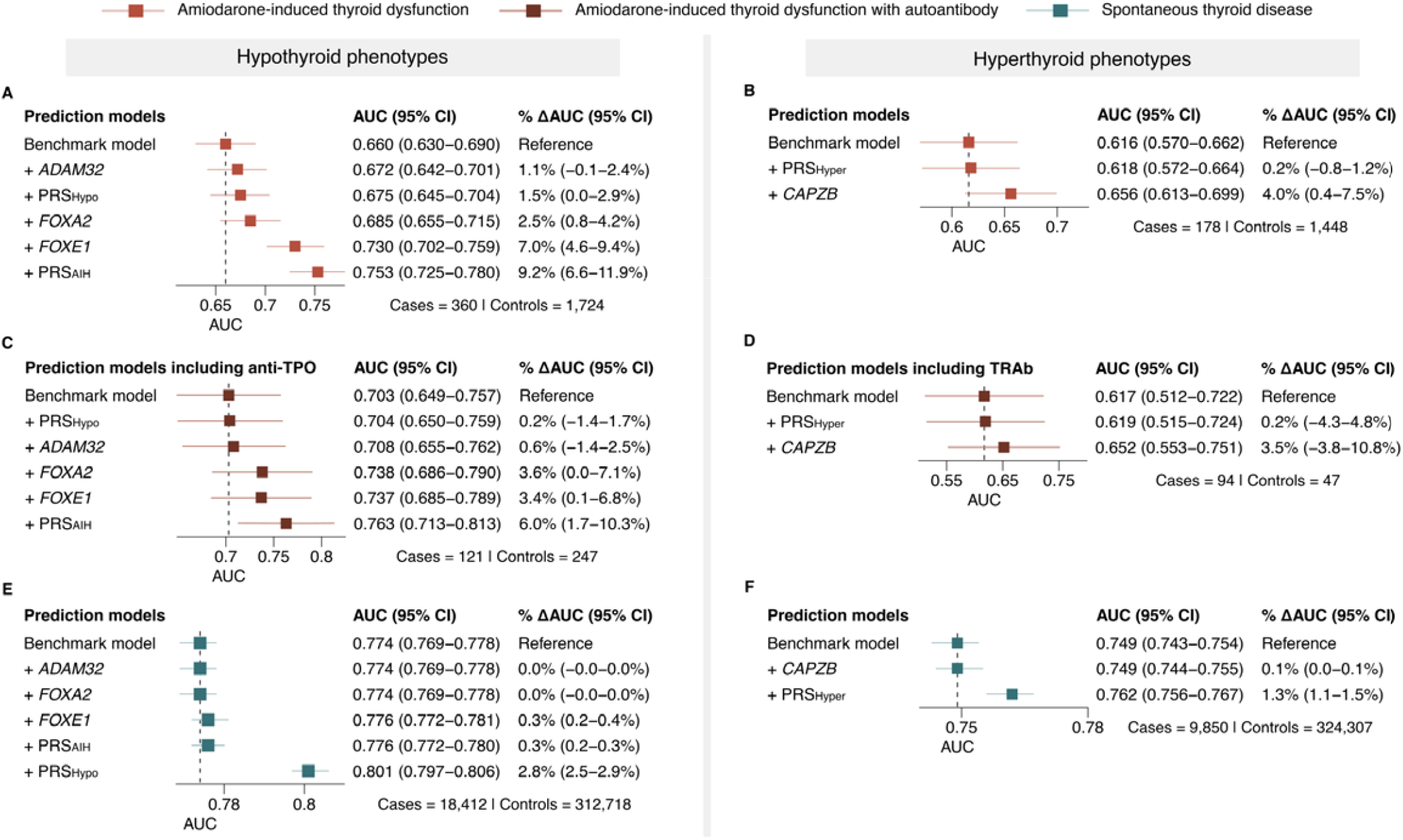
Genetic predictors of amiodarone-induced thyroid dysfunction compared to spontaneous thyroid disease. The panels illustrate the change in predictive power (ΔAUC) after adding genetic risk factors to a benchmark model (age, sex, pre-existing thyroid disease and four principal components). Evaluated genetic factors were amiodarone-induced thyroid dysfunction risk alleles, PRS_AIH_, and genome-wide PRSs for spontaneous hypo-or hyperthyroidism. Models were used to predict: (**A**) amiodarone-induced hypothyroidism (AIH), (**B**) amiodarone-induced thyrotoxicosis (AIT), (**C**) AIH in individuals with anti-TPO, (D) AIT in individuals with TRAb, (**E**) spontaneous hypothyroidism or (**F**) spontaneous hyperthyroidism. Abbreviations: AUC, area under the curve; PRS, polygenic risk score.

### Clinical application of genetic screening using ADR-associated variants

To evaluate the clinical utility of these variants, we assessed their potential for genotype-guided risk stratification. For AIH, screening for two risk alleles of the identified variants in *FOXE1* and *FOXA2* yielded NPVs of approximately 95% with PPVs ranging from 13–23% (in CHB/DBDS, deCODE genetics and with external validation in UKB primary care data; **Table 2**). In lower deciles of the PRS_AIH_, genetic screening could also identify individuals at low risk of AIH (i.e., NPV ranging from 92.1–98.6% in the lower 20^th^ percentile with PPVs from 19.5–22.2%; **Table 2** and **Supplementary Table 13**). The number needed to genotype (NNG) to capture one AIH case using a PRS_AIH_ cut-off in the lower 20^th^ percentile as a positive test was 26 in CHB/DBDS and 30 in UKB. For AIT, screening for two risk alleles of the *CAPZB* variant yielded NPVs ranging from 91.9–97.5% and PPVs of 1.9–13.9% (**Table 2**). Consistent with the low population prevalence of AIT, the associated NNG values were substantially higher: 106 in CHB and 5,155 in UKB.

**Table 2.** Predictive values of genetic screening for amiodarone-induced thyroid dysfunction associated variants. Positive and negative predictive values for screening of either risk alleles of the AIH-associated variants, a PRS_AIH_, or the AIT-associated variant. Predictive values are shown for the Copenhagen Hospital Biobank and Danish Blood Donor Study (CHB/DBDS), deCODE genetics, and UK Biobank primary care data (used as an external validation cohort and was not included for discovery of the genetic risk factors for amiodarone-induced thyroid dysfunction). AIH sample sizes were: CHB/DBDS, 360 cases and 1,724 controls; deCODE genetics, 248 cases and 1,006 controls; UK Biobank primary care data, 20 cases and 484 controls. AIT samples sizes were: CHB/DBDS, 178 cases and 1,448 controls; deCODE genetics 36 cases and 1,419 controls; UK Biobank primary care data, 49 cases and 403 controls. Abbreviations: AIH, amiodarone-induced hypothyroidism; AIT, amiodarone-induced thyrotoxicosis; PPV, positive predictive value; NPV, negative predictive value.

| Gene | Test | Condition | CHB/DBDS |  | deCODE genetics |  | UKB primary care |  |
| --- | --- | --- | --- | --- | --- | --- | --- | --- |
|  |  |  | PPV (95%CI) | NPV (95%CI) | PPV (95%CI) | NPV (95%CI) | PPV (95%CI) | NPV (95%CI) |
| *FOXE1* | Two alleles<br>vs. rest | AIH | 26.1%<br>(24.4 – 27.8) | 90.4%<br>(88.9 – 91.8) | 32.5%<br>(29.7 – 35.6) | 87.8%<br>(86 – 89.4) | 18.1%<br>(15 – 21.6) | 94.3%<br>(91.5 - 96.2) |
| *FOXA2* | Two alleles<br>vs. rest | AIH | 21.4%<br>(19.9 – 22.9) | 87.2%<br>(85.4 – 88.7%) | 24.2%<br>(22.2 – 26.2) | 85.3%<br>(82.8 – 87.4) | 12.2%<br>(9.6 – 15.3) | 90.5%<br>(87.2 – 93.1) |
| *FOXE1+FOXA2* | > Two alleles<br>vs. rest | AIH | 18.5%<br>(18.1 – 18.8) | 94.3%<br>(90.2 – 96.8) | 21.3%<br>(20.9 – 21.8) | 95.0%<br>(89.4 – 97.7) | 12.1%<br>(11.5 – 12.7) | 98.1%<br>(88.2 – 99.7) |
| $PRS_{AIH}$ | Bottom 20%<br>vs. rest | AIH | 19.5%<br>(17.7 – 21.5) | 94.4%<br>(91.4 – 96.6) | 22.7%<br>(20.2 – 25.4) | 92.9%<br>(88.8 – 95.8) | 12.8%<br>(9.6 – 16.6) | 98.7%<br>(93.0 – 100.0) |
| *CAPZB* | Two alleles<br>vs. rest | AIT | 13.9%<br>(12.5 – 15.5) | 91.9%<br>(90.3 – 93.3) | 2.6%<br>(1.0 – 6.4) | 97.5%<br>(97.3 – 97.8) | 1.9%<br>(0.3 – 11.9) | 95.8%<br>(95.4 - 96.2) |

## DISCUSSION

We identified four variants that were significantly associated with amiodarone-induced thyroid dysfunction. Three of these variants mapped to genes with established roles in thyroid development, pathophysiology, and hormone levels: *FOXE1, FOXA2*, and *CAPZB*.^34–36^ The fourth finding at *ADAM32* should be interpreted with caution. The variant lacked convincing evidence from prior studies or functional analyses and would require further investigation to establish a potential biological relevance. Further, while the three variants were previously associated to thyroid disease and hormones, we showed that their effect sizes were significantly larger for amiodarone-induced thyroid dysfunction. Collectively, our findings suggest a distinct genetic architecture and that these loci confer a specific susceptibility to iodine-induced thyroid stress, unmasked in this case by amiodarone.

The pathogenesis of amiodarone-induced thyroid dysfunction likely centers on two factors: the high iodine load from amiodarone therapy, and the thyroid’s autoregulatory response. Amiodarone is 37% iodine by weight^37^ and a standard 200 mg dose releases 50 - 100 times the daily recommended iodine intake.^2^ This iodine excess triggers the autoregulatory ‘Wolf-Chaikoff’ effect which inhibits thyroid hormone synthesis.^37^ We suggest that genetically susceptible individuals cannot maintain this autoregulation, leading to thyroid dysfunction through one of two pathways: 1) failure to escape the Wolf-Chaikoff effect, causing hypothyroidism, or 2) disrupted autoregulation resulting in thyrotoxicosis.^38^ Our findings point to specific biological pathways involvement: *FOXE1* is critical for thyroid autoregulation, as it regulates transcription of genes essential for hormone synthesis and transport, including *NIS, TPO*, and *TG*.^39^ Carriers of the *FOXE1* risk variant may have impaired autoregulation, failing to escape the Wolf-Chaikoff effect, and thus becoming susceptible to hypothyroidism. The *FOXA2-*association may involve dysregulation of the hypothalamic-pituitary-thyroid (HPT) axis, as loss-of-function mutations in *FOXA2* are linked to a syndrome including central hypothyroidism.^40^ We know that common GWAS signals often converge with Mendelian disorder loci,^41^ and hypothesize that this variant impairs HPT axis function, an effect likely originating in the thyroid. This is supported by our findings that the variant is both a thyroid eQTL for *FOXA2* associated with *trans*-pQTLs consistent with a hypothyroid phenotype (increased TSH and reduced fT4 and Tg levels). Finally, *CAPZB* encodes a protein involved in the TSH-mediated endocytosis of thyroid colloid, a crucial step for hormone transport.^42^ The *CAPZB* variant was also a thyroid eQTL for *CAPZB*, likely disrupting hormone autoregulation during iodine excess. This mechanism would explain the variant’s association with AIT and the observed downstream effects on reduced TSH and increased fT4 levels.

The current risk assessment for amiodarone-induced thyroid dysfunction is reactive and with variable predictive reliability. Our findings provide a new proactive solution to better stratify risk pre-therapy. We demonstrated that common variants in *FOXE1, FOXA2*, and *CAPZB* are superior predictors of amiodarone-induced thyroid dysfunction which offered significant independent information beyond age, sex, genome-wide PRSs, and to some extent thyroid autoantibodies. The primary clinical strength of genetic screening lies in the very high NPV ≈ 95% which can immediately de-risk the estimated 15% of the population who lacks the risk alleles. This is a clinically actionable outcome, providing reassurance to patients and physicians, and a rationale to support more confident amiodarone dosing and less resource-intensive monitoring. The clinical reassurance benefits are especially important in AIT, which is associated with high morbidity and mortality.^4^ As genotyping costs decrease and preemptive genetic testing becomes more integrated in routine care, testing for these variants could help improve the safe and effective use of amiodarone.

This study has several limitations. First, our findings will require validation in larger, independent cohorts. Second, our reliance on diagnostic codes from electronic health records introduces a risk of phenotype misclassification. Third, the AUCs were calculated without external validation and are therefore subject to potential overfitting, and the autoantibody sub-analysis was limited by sampling bias, as tests were performed based on clinical indication, warranting cautious interpretation. Further work is needed to investigate mechanisms by which the variants affect iodine handling and autoregulation, as these roles may extend beyond amiodarone-induced thyroid dysfunction. In conclusion, our study provides evidence that genetic variants with high frequency and effect in thyroid genes influence risk of amiodarone-induced thyroid dysfunction. The high NPVs of genotype-based screening highlights the potential to identify patients at low risk which could allow for personalized monitoring and treatment.

## Data Availability

Genome-wide summary statistics will be made available to the public on the GWAS Catalog.

## ACKNOWLEDGMENTS

This research was conducted using the UKB resource under application number 43247 and 56270. This work was supported by BRIDGE - Translational Excellence Programme (nos. NNF18SA0034956 and NNF20SA0064340), The Beckett Foundation, Agnes and Knut Mørks Foundation, The Innovation Fund Denmark (PM Heart), NordForsk, Villadsen Family Foundation, The Arvid Nilsson Foundation, Sygeforsikringen “danmark”, Estonian Research Council grants PRG1911, the Estonian Center of Genomics/Roadmap II (TT17), and the Ministry of Education and Research Centers of Excellence grant TK214 (Centre of Excellence for Personalized Medicine), and Novo Nordisk Foundation (nos. NNF17OC0027594 and NNF14CC0001). J.G. is supported by AUFF Recruitment grant (AUFF-E-2024-7-10). All human research was approved within each contributing study by the relevant institutional review board (CHB and DBDS: National Committee on Health Research Ethics; deCODE genetics: National Bioethics Committee (VSN-15-057); UKB: Northwest Multicenter Research Ethics Committee; Estonian Biobank: Estonian Committee on Bioethics and Human Research) and conducted according to the Declaration of Helsinki. All participants provided written informed consent, except for CHB, where patients were informed about the opt-out possibility of having their biological specimens excluded from use in research. Since 2004, a national Register on Tissue Application (Vævsanvendelsesregistret) lists all individuals who have chosen to opt out and whose samples cannot be used for research purposes. Before initiating this study, individuals listed in the Register on Tissue Application were excluded.

## CONTRIBUTIONS

S.A.R., J.B., G.A., H.B., J.G., V.T., A.H., and S.S. conceived the study. S.A.R., J.G., V.T., S.R., S.S., A. H., D.F.G., M.T.L., S.K., and M.K. performed analyses in the respective datasets. J.G., H.B., S.R.O., O.B.P., S.B., S.S., I.J., D.F.G., G.H., E.F., H.H., A.H., K.S., M.T.L., A.M.S., W.H., P.N., and A.M. supervised analyses in their respective datasets. S.A.R., J.B., G.A., J.G., H.B., S.R.O., O.B.P., S.S.,V.T., S.R., A.H., S.S., D.F.G, M.K., and M.T.L contributed to writing the manuscript. S.A.R., J.B, J.G., S.R., and V.T. performed meta-analysis and created figures and tables. S.A.R., J.B., G.A., J.G., H.B., V.T., S.R., G.H., I.J., E.F., H.H., S.S., A.H., D.F.G., M.K., S.K., W.H., A.M.S., and M.T.L. contributed to downstream analysis and drafted the manuscript. S.A.R, J.B., G.A., M.S., C.E., M.T.B., B.A., H.U., S.B., S.R.O., N.B., J.T., C.M., B.D.M., O.B.P, E.S., S.S., H.B., J.G., V.T., S.R., G.H., I.J., E.F., H.H., S.S., A.H., D.F.G, K.S., P.N., M.K., and M.T.L interpreted the results and reviewed and commented on the manuscript.

