## Supplementary Information for "GWAS of amiodarone-induced thyroid dysfunction: Applications for genotype-guided risk stratification"

*By Rand S. et al.*

**Table of content**

**Supplementary Note**

*Case-control definitions used in the study*	Page 2

*Copenhagen Hospital Biobank and The Danish Blood Donor Study*	Page 2-3

*deCODE genetics*	Page 3-4

*The Estonian Biobank*	Page 4

*Mass General Brigham Biobank*	Page 4-5

*The UK Biobank*	Page 5

**Supplementary Figures**

*Supplementary Figures 1A-D*	Page 6

*Supplementary Figures 2A-D*	Page 7

*Supplementary Figure 3A-H*	Page 8

**References**	Page 9

**SUPPLEMENTARY NOTE**

**Case and control definitions.**

We applied a uniform criterion across all datasets to define amiodarone-induced hypothyroidism (AIH) and amiodarone-induced thyrotoxicosis (AIT).

**AIH.** Individuals were first identified as amiodarone users through the Anatomical Therapeutic Chemical (ATC) classification code *C01BD01*. To capture hypothyroidism most likely related to amiodarone, we considered events occurring within 365 days of treatment initiation, based on prior evidence.^1^ Cases were defined using ICD-10 codes *E032* (“Drug-induced hypothyroidism”), *E038* (“Other specified hypothyroidism”), *E039* (“Hypothyroidism, unspecified”) or ATC code *H03A* (thyroid hormone replacement therapy). Controls were defined as amiodarone users who:

1. had no diagnosis of hypothyroidism and were never prescribed thyroid hormone therapy before, during, or after amiodarone use, and
2. had at least one-year of follow up and three amiodarone prescriptions within one year.

**AIT.** Amiodarone users were identified by ATC code *C01BD01*. Thyrotoxicosis events were assessed within 550 days of treatment initiation, in line with previous studies.^1^ Cases were defined using ICD-10 codes *E058* (“Other specified thyrotoxicosis”) and *E059* (“Thyrotoxicosis, unspecified”) or ATC code *H03B* (anti-thyroid drugs). Controls were defined as amiodarone users who:

1. had no diagnosis of thyrotoxicosis and were never prescribed anti-thyroid therapy before, during, or after amiodarone use, and
2. had at least one-year of follow up and three amiodarone prescriptions within one year.

**Cohort information on ethics and genotyping and imputation details**

**Copenhagen Hospital Biobank (CHB) and The Danish Blood Donor Study (DBDS).**

Copenhagen Hospital Biobank (CHB) is a hospital-based biobank that includes nearly 360,000 patients, and for the current study, we undertook analysis in The Oral Cardiometabolic Health Study (CHB-OCMS), a sub-cohort of CHB. Patient samples were collected under admissions to general hospitals in The Capital Region of Denmark (Region Hovedstaden) between 2009 and 2020 and genotyped with samples collected from DBDS.^2^ DBDS is a nation-wide cohort study including over 175,000 blood donors, of which 116,000 were genome-wide genotyped and used in this study.^3^ Both studies (CHB/DBDS) link patients’ genetic profiles with extensive and longitudinal electronic health data from Danish national registries.

Genotyping and imputation: CHB and DBDS participants were genotyped with Illumina Global Screening Array (GSA). Sample-level quality control (QC) excluded individuals with call rate < 97%, ambiguous sex, genetic ancestry outliers or heterozygosity outliers > 5 SD from the mean. Variant-level QC excluded single nucleotide polymorphisms (SNPs) with call rate < 98%, low Hardy-Weinberg Equilibrium (HWE) *P* < 1 × 10^-5^, or minor allele frequency (MAF) < 0.1%. Genotyped data were phased and imputed using whole-genome sequencing from 50,179 European ancestry individuals.^4^

Ethics: The biological samples stored in CHB-OCMS were residual material from blood sampling, and therefore participants were informed about an opt-out possibility. Participants from DBDS provided informed consent. In Denmark the national Register on Tissue Application (Vævsanvendelsesregistret) since 2004 lists all individuals who have chosen to opt out and whose samples cannot be used for research purposes. Individuals that opted out were excluded from all analyses. CHB-OCMS and DBDS are approved by the Regional and National Committee on Health Research Ethics (SJ-989 and NVC 1700407) and the Danish Data Protection Agency (P-2022-913 and P-2019-99).

**deCODE genetics**

deCODE genetics is an Icelandic study population genotyped by deCODE genetics/Amgen, a biopharmaceutical company, and includes genome information on more than two-thirds of the adult population of Iceland.

Genotyping and imputation: In deCODE genetics, 173,025 Icelanders were genotyped on multiple Illumina platforms and imputed against a reference panel based on whole-genome sequencing of 63,460 Icelanders using Illumina TruSeq methodology (mean depth 39.8×; SD 14.2; range: 20.0× – 397.8×).^5,6^ Long-range phasing and imputation extended coverage to first- and second-degree relatives using genealogic data, without storing raw genotypes. SNPs and indels were called in CHB, DBDS, and deCODE genetics using GraphTyper (v.2.7.1).^7^

Ethics: The study was approved by the Icelandic Data Protection Authority and the National Bioethics Committee (VSN-16-042, VSN-15-023). All participants that included blood samples for research purposes provided informed consent. In accordance with the regulations by the Icelandic Data Protection Authority sample identifiers were encrypted.

**The Estonian Biobank (EstBB).**

The EstBB is a population-based biobank established in 2000 that holds genetic data for more than 213,000 inhabitants of Estonia (20% of the adult population). Participants were were recruited nationwide through general practitioners and dedicated recruitment offices. At baseline, participants underwent a standardized health examination, provided blood samples and completed questionnaires covering lifestyle, diet, and clinical diagnoses. The database is regularly updated through linkage with national electronic databases and registries.^8^

Estonian Biobank research team refers to Andres Metspalu, Lili Milani, Tõnu Esko, Reedik Mägi, Mait Metspalu, Mari Nelis, and Georgi Hudjashov, giving them credit for data collection, genotyping, quality control and imputation.

Genotyping and imputation: EstBB participants were genotyped on Illumina GSA. Sample-level QC excluded individuals with call rate < 95% or sex discordance between genotype and phenotype. Variant-level QC excluded SNPs with call rate <95%, MAF <1%, or extreme HWE deviation (*P* < 1 × 10^−4^). Imputation was performed using a population-specific haplotype reference panel based on whole-genome sequencing data from 2,056 Estonians.

Ethics: The EstBB project is being conducted according to the Estonian Human Genes Research Act, and all participants have signed a broad informed consent form. This study was approved by the Estonian Committee on Bioethics and Human Research, Estonian Ministry of Social Affairs (no. 1.1-12/624 approved on 24 March 2020).

**Mass General Brigham Biobank.**

The Mass General Brigham Biobank (MGBB; formerly Partners HealthCare Biobank) is a large-scale biobanked linked with extensive electronic health record (EHR) data and survey information, enabling investigations of genomic, environmental, biomarker, and family history associations with disease phenotypes. To date, MGBB has enrolled more than 135,000 participants and generated genomic data for over 65,000 individuals.^9^

Ethics: Informed consent was obtained from all participants in MGBB. The study was conducted in accordance with the Declaration of Helsinki and approved by the Institutional Review Board (or Ethics Committee) of Mass General Brigham (protocol code 2009-P-002312 approved on 29 April 2010).

Genotyping and imputation: MGBB participants were genotyped on Illumina GSA. Sample-level QC removed individuals with call rate < 98% or ambiguous sex, while variant-level QC excluded SNPs with call rate < 98%, minor allele count ≤ 2, or HWE *P* < 1 × 10^-6^. Phasing and imputation were performed with Minimac3 using the Haplotype Reference Consortium Panel 1.1 as reference.

Ethics: Informed consent was obtained from all participants in MGBB. The study was conducted in accordance with the Declaration of Helsinki and approved by the Institutional Review Board (or Ethics Committee) of Mass General Brigham (protocol code 2009-P-002312 approved on 29 April 2010).

**UK Biobank**

The UK Biobank is a longitudinal cohort study with including around 500,000 volunteering residents of the United Kingdom. Participants were between 40 and 69 years of age at time of recruitment (2006-2010).^10^ In the present study, we used the UK Biobank primary care data covering 245,000 participants (available for non-Covid research purposes). The UKB data resource was accessed under Application ID 43247 and 56270.

Ethics: All study participants provided written informed consent to participate in the study, and the project were ethically approved by the Northwest Multicenter Research Ethics Committee, UK (Ref: 16/NW/0274).

**SUPPLEMENTARY FIGURES.**

**Supplementary Figure 1. Regional associations plot for genome-wide association to AIT and AIH.** The x-axis depicts the genomic region of the lead variant, while the y-axis shows the strength of association to the phenotype (-log_10_(*P*)). Dots are colored according to their linkage disequilibrium with each lead variant. The bottom panels include protein coding genes in the genomic region, where exons are colored dense orange. The red dotted line marks the threshold for genome-wide significance. **A.** Regional associations for the chromosome 1 AIT-lead variant rs867355. **B.** Regional associations for the chromosome 8 AIH-lead variant rs1268751. **C.** Regional associations for the chromosome 9 AIH-surrogate lead variant rs1443438 (linkage disequilibrium with actual lead variant rs36052460 R^2^ = 1, which was unavailable in the LD reference and data used to create the plots) . **D.** Regional associations for the chromosome 20 AIH-lead variant rs2424459.

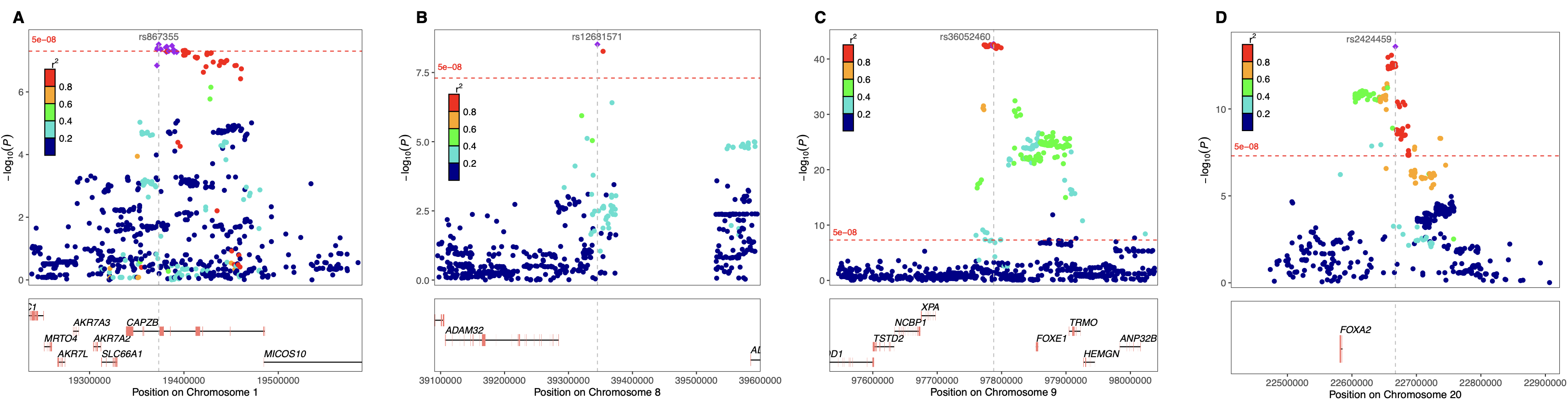

**Supplementary Figure 2. Colocalization in regions of lead variants for amiodarone-induced thyroid disease and their counterpart spontaneous thyroid disease.** Genome-wide associations for AIT and AIH were queried in published GWAS for hypothyroidism and hyperthyroidism. *P*-values for variants were then compared between the amiodarone-induced thyroid disease and its spontaneous thyroid disease counterpart. Variants are colored according to their linkage disequilibrium with the lead variant in the locus. **A.** *CAPZB* locus for AIT and hyperthyroidism*.* **B.** *ADAM32* locus for AIH and hypothyroidism*.* **C.** *FOXE1* locus for AIH and hypothyroidism. **D.** *FOXA2* locus for AIH and hyperthyroidism.

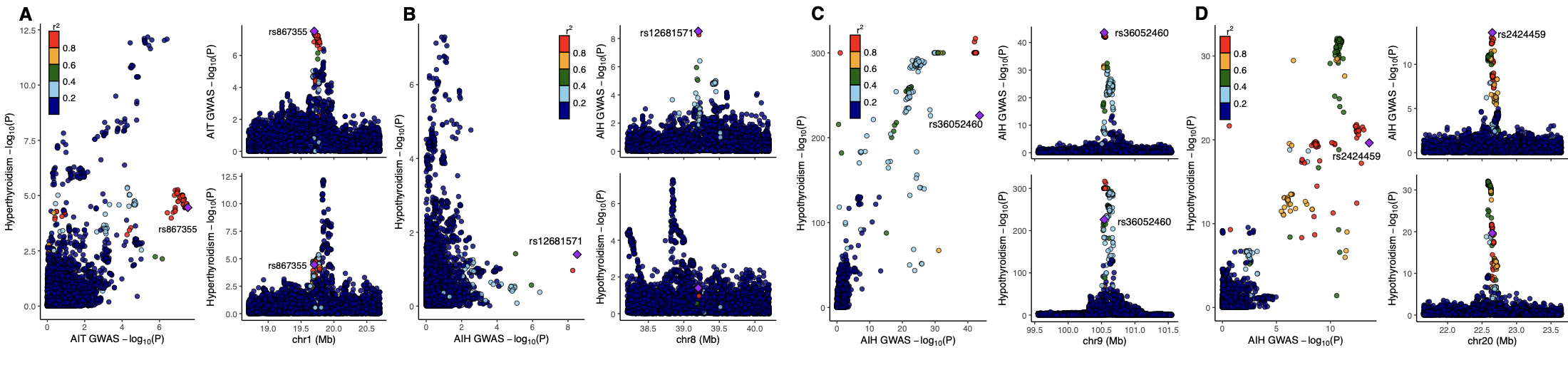

**Supplementary Figure 3. Mapping of expression quantitative loci (eQTLs) for genes in the vicinity of amiodarone-induced thyroxicosis lead variant rs867355. A-D.** The AIH lead variant rs36052460 was unavailable in eQTL data, so we instead used the next top variant to extract eQTL associations (r2 between the two = 0.999)**.** Regional association plots for the rs1443438 for associations in the thyroid and for *FOXE1*-associations. **E-F.** Regional association plots for the rs2424459 variant in thyroid tissue. **G-H.** Regional association plots *CAPZB* and neighboring gene *PQLC2* in tissues thyroid tissue.

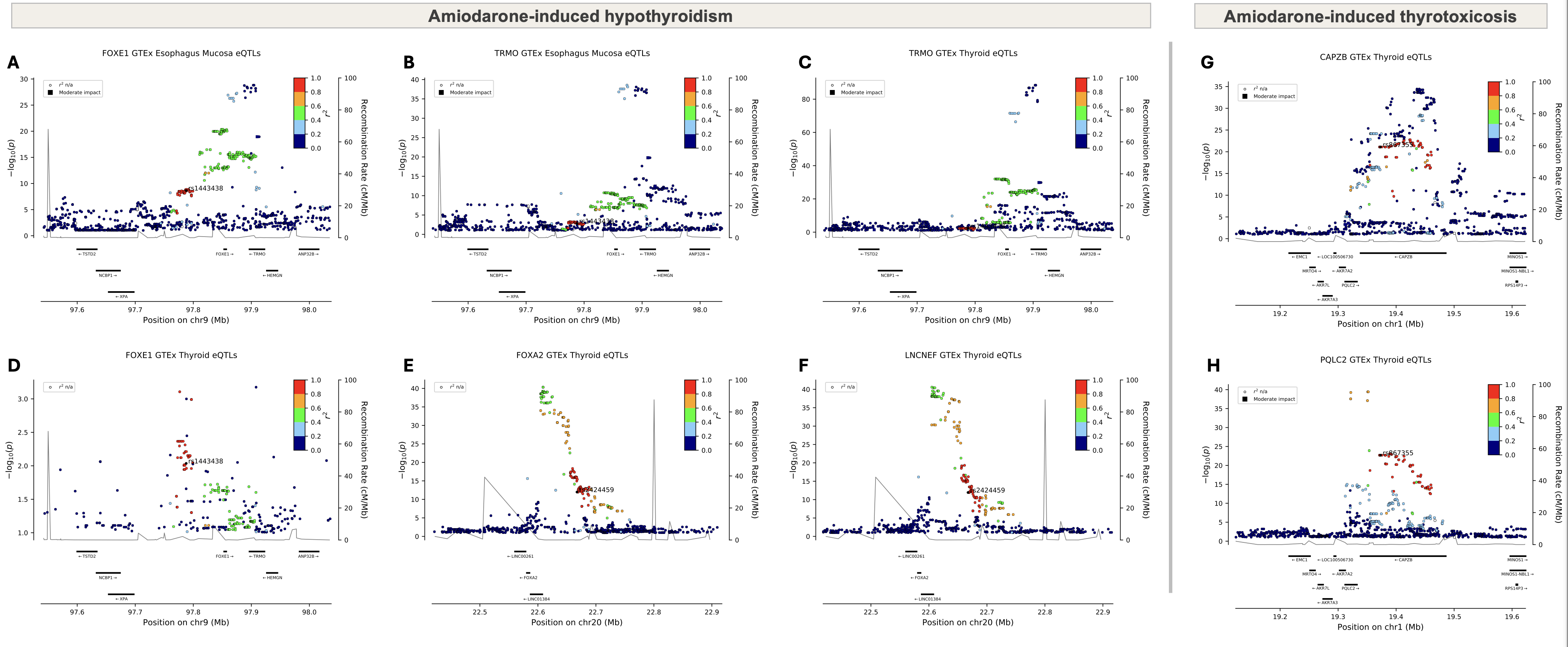
